# Diet Explains Significant Variance in Oral Microbial Community Structure

**DOI:** 10.64898/2026.04.24.26351661

**Authors:** Yu Xie, Mengning Bi, Wen Gu, Yuan Li, Andrea Roccuzzo, Bob T. Rosier, Maurizio S. Tonetti

## Abstract

Diet is an important ecological modulator of the oral microbiome, yet population-level evidence on a broader spectrum of food components remains limited. This cross-sectional study investigated associations among dietary intake, oral rinse microbiome, and oral disease conditions in a nationally representative sample of United States adults from the National Health and Nutrition Examination Survey. A total of 3,254 participants with oral rinse microbiome sequencing data were included, with oral conditions classified as oral health, caries-only, periodontitis-only, or co-existing disease. Dietary intake was assessed using 24-hour dietary recalls and summarized as dietary indices and energy-adjusted food components. Associations between diet and the oral microbiome were evaluated using community-level analyses, regression models, mediation analyses, and unsupervised clustering, while accounting for oral conditions. This study found that dietary intake, as a combined variable set, explained 3.6% of the variance in oral rinse microbial community structure; this was comparable to oral disease status or smoking and larger than sociodemographic factors. Healthier dietary profiles, including higher health-associated dietary index scores and greater vegetable and fruit intake, were associated with taxa commonly linked to oral health (e.g., *Neisseria, Cardiobacterium and Lautropia*). In contrast, added sugars, alcoholic drinks, cured meat, potatoes, dairy products, and higher dietary inflammatory index scores showed opposite association patterns. Mediation analyses suggested that coordinated microbial groups may partly link dietary exposures with oral disease outcomes, particularly for vegetables and added sugars. Additionally, three population-level dietary patterns were identified, among which the plant-rich pattern was associated with more favorable oral health and microbial profiles enriched in nitrate-reducing commensals, including *Neisseria* and *Haemophilus*. Overall, dietary intake was associated with oral microbiota composition and oral health conditions, supporting ecological influences of dietary components beyond sugar on oral bacteria and dental diseases. Longitudinal studies are needed to clarify the direction and causality of these relationships.

## Introduction

The oral cavity is a dynamic and accessible microbial ecosystem. Its microbiome occupies multiple oral niches and is shaped by continuous interactions among microorganisms, host factors, and the local environment (Sedghi et al. 2021). In oral health, these communities exist in a stable, balanced state (i.e., normobiosis), maintained by salivary factors (e.g., pH buffering and glycoproteins as microbial nutrients), host defenses, and microbial interactions (Rosier et al. 2018). When this balance is disrupted by changes in host response, local or systemic conditions, dietary exposures, or poor oral hygiene, the microbiota may shift from normobiosis to dysbiosis (i.e., an increase in disease-associated species and functions) (Santonocito et al. 2022). Such ecological disruption contributes to dental caries and periodontitis, and has also been linked to systemic conditions (Sedghi et al. 2021).

The Ecological Plaque Hypothesis proposes that changes in nutrient availability and local physicochemical conditions can drive transitions from health-associated communities to dysbiotic states (Marsh 2003). Within this framework, dietary intake is considered an important ecological determinant of the oral microbiome. The clearest example is the relationship between fermentable carbohydrates and dental caries, in which frequent exposure promotes acid production, lowers biofilm pH, and favors acidogenic and acid-tolerant microorganisms, which can ultimately lead to enamel demineralization and dental caries (Moores et al. 2022). Historically, following the development of agriculture and the Industrial Revolution, increased carbohydrate consumption promoted the expansion of cariogenic bacteria in the oral cavity and may have contributed to the emergence of dental caries (Adler et al. 2013). Interestingly, one study reported that gingival inflammation did not increase despite the absence of oral hygiene when participants consumed a diet low in processed carbohydrates, suggesting a potential link between carbohydrate intake and periodontal disease (Baumgartner et al. 2009).

More broadly, diet may influence oral microbial ecology by supplying substrates, altering redox balance, and modulating metabolic interactions and pH within biofilms. Emerging evidence suggests that nitrate, a plant-derived compound abundant in certain vegetables, may support nitrate-reducing bacteria and help maintain oral microbiome equilibrium (Jockel-Schneider et al. 2021; Rosier et al. 2022). In contrast, diets with higher inflammatory potential and higher Dietary Inflammatory Index (DII) value (e.g., low in plant-derived components and high in sugars, saturated, and trans fats) have been associated with poorer systemic and oral health (Grosso et al. 2022; Li et al. 2021), and may contribute to microbiome dysbiosis through host inflammatory responses and ecological changes (Santonocito et al. 2022).

Although these mechanisms are biologically plausible, several important knowledge gaps remain. Most studies have focused on isolated nutrients or selected food components, such as carbohydrates, fats, proteins, or nitrate, whereas real-world diets consist of complex food combinations containing diverse nutrients (i.e., the food matrix). These combinations may produce effects on the oral microbiome that differ from or even oppose those of the individual components. For example, although beetroots are rich in sugars, their consumption has been associated with increased oral pH, likely due to other components (e.g., nitrate and antioxidants), contrasting with the typical acidification observed after sugar intake (Burleigh et al. 2020). Similarly, consuming nitrate-rich vegetables prior to sugar intake can limit oral acidification. In addition, the relative contributions of dietary intake, health status, and other sociodemographic factors to oral microbial variation remain poorly characterized at the population level. Another challenge is that dental caries and periodontitis themselves strongly affect microbial composition, making it difficult to distinguish dietary effects from disease-related changes (Sedghi et al. 2021; Xie et al. 2026). Finally, although mediation analyses involving the microbiome have been explored, most studies have focused on individual taxa or diversity indices (Nath et al. 2025; Yue et al. 2025a), and whether coordinated microbial communities mediate diet-disease associations remains unclear.

These questions are particularly suited to oral rinse microbiome data in population-based studies. Oral rinse samples provide a practical whole-mouth representation of oral microbial communities and capture a broader ecological snapshot than site-specific plaque sampling, while remaining feasible in large epidemiologic settings (Xie et al. 2026).

Against this background, we used oral rinse microbiome data from the National Health and Nutrition Examination Survey (NHANES) program to investigate diet-microbiome associations in a nationally representative adult population. As outlined in **Figure 1A**, the aims of this study were: (i) to examine associations between oral rinse microbiota and a broad range of dietary intake variables while accounting for oral conditions, to identify diet-microbiome associations independent of oral disease status; (ii) to assess whether coordinated microbial community groups mediate associations between dietary intake and oral conditions; and (iii) to derive real-world dietary patterns using unsupervised clustering and evaluate their associations with oral disease distributions and corresponding oral rinse microbial community features.

**Figure 1.**
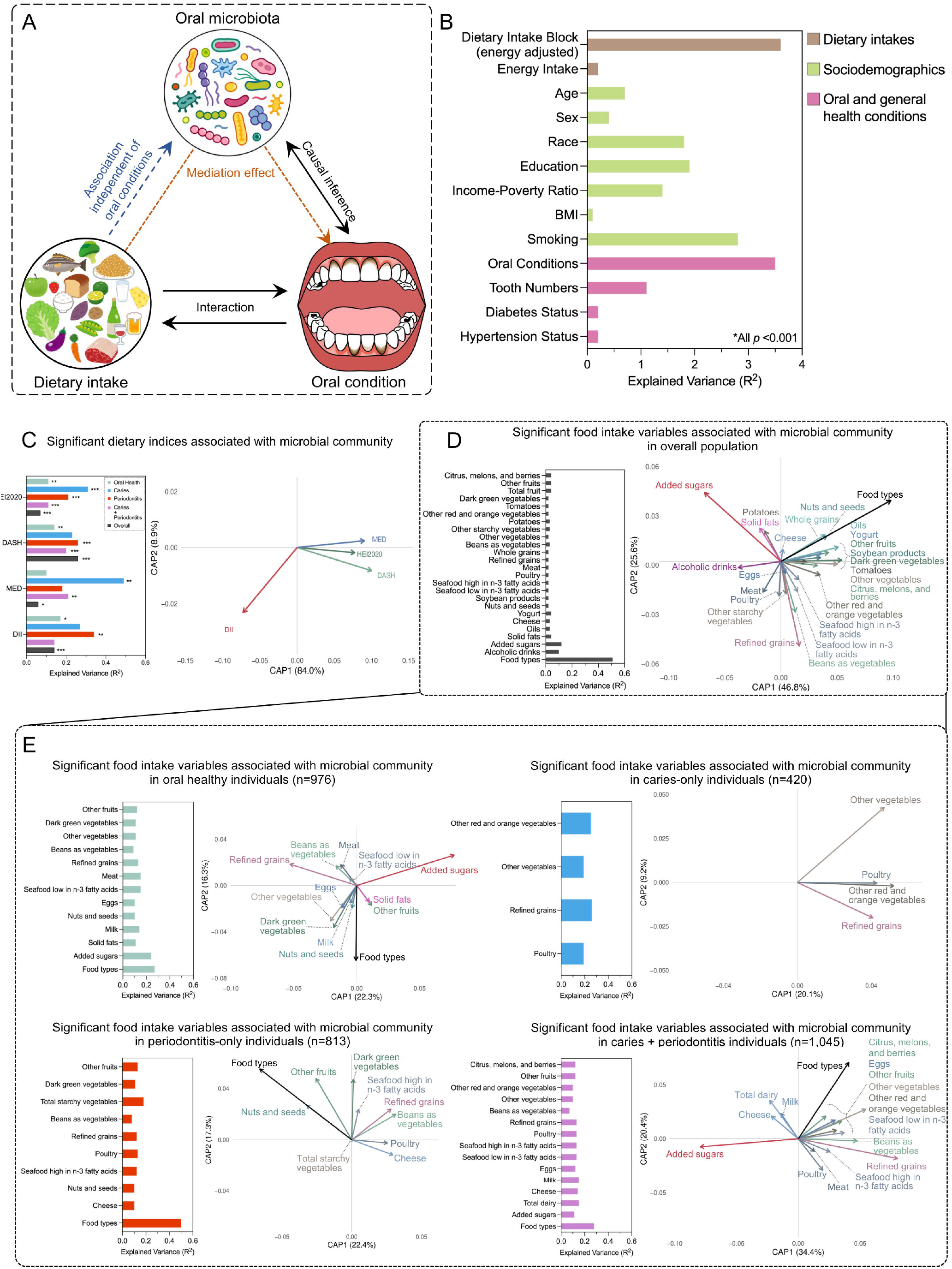
Dietary intake explains variation in the overall oral rinse microbial community structure. (A) Study diagram. To evaluate the associations between dietary intake and oral rinse microbiota independent of oral conditions, and further assess whether oral rinse microbiota mediates the association between dietary intake and oral conditions; (B) Variance in microbial community structure based on Aitchison dissimilarities explained by dietary intake, sociodemographic factors, and oral and general health conditions. Explained variance (R^2^) and *p* values were calculated using PERMANOVA. The dietary intake block represents energy-adjusted food intake variables, which were simultaneously included in a multivariable PERMANOVA model to estimate the cumulative variance explained by dietary intake. For other factors, R^2^ was evaluated using single-variable PERMANOVA models. (C) Distance-based redundancy analysis (dbRDA) under Aitchison distance showing significant dietary indices associated with variation in the overall oral rinse microbial community in the total population. The corresponding explained variance (R^2^) in the total population and within each oral condition subgroup is summarized in the bar plot; (D) Significant energy-adjusted food intake variables associated with overall microbial community variation in the total population, identified by dbRDA under Aitchison distance. The corresponding R^2^ is presented in the bar plot; (E) Significant energy-adjusted food intake variables associated with microbial community variation within different oral conditions of oral health, caries-only, periodontitis-only, and co-existing caries and periodontitis groups, identified by dbRDA under Aitchison distance. The corresponding R^2^ is presented in the bar plot. For dbRDA plots in panels (C-E), arrows represent dietary indices or food intake variables significantly associated with microbial community structure. Arrow length reflects the strength of association with the constrained ordination axes. The absolute direction of an arrow is arbitrary; however, the relative angles between arrows indicate whether variables are associated with the microbial community in a similar or contrasting direction in the constrained ordination space. Variables with relative angles < 90° show associations in a similar direction, whereas variables with relative angles > 90° indicate associations in opposing directions. In panel (C), statistical significance is denoted as *: *p* <0.05, **: *p* <0.01, ***: *p* <0.001. In panels (D) and (E), food intake variables were adjusted for total energy intake and expressed per 1,000 kcal in analyses. During all analyses, statistical significance was determined using Benjamini-Hochberg false discovery rate (FDR) correction. Abbreviations in figure: HEI2020 = Healthy Eating Index 2020; DASH = Dietary Approaches to Stop Hypertension; MED = Mediterranean Diet score; DII = Dietary Inflammatory Index; CAP = canonical axis in constrained ordination.

## Methods

Details of the methodology are provided in the Appendix Methods, including comprehensive descriptions of the study design and population, clinical examinations, dietary intake assessments, oral rinse sample collection, sequencing and data processing procedures, and bioinformatic and statistical analyses. The article complies with the STROBE checklist.

## Results

A total of 3,254 participants from NHANES with 77 genus-level taxa were included in this study **(Appendix Figure 1)**. The participants were divided among different oral conditions: 976 were defined as oral health, 420 as caries-only, 813 as periodontitis-only, and 1,045 as co-existing caries and periodontitis. Sociodemographic, behavioral, and systemic characteristics differed significantly across oral condition groups, except energy intake **(Table 1)**.

**Table 1.**
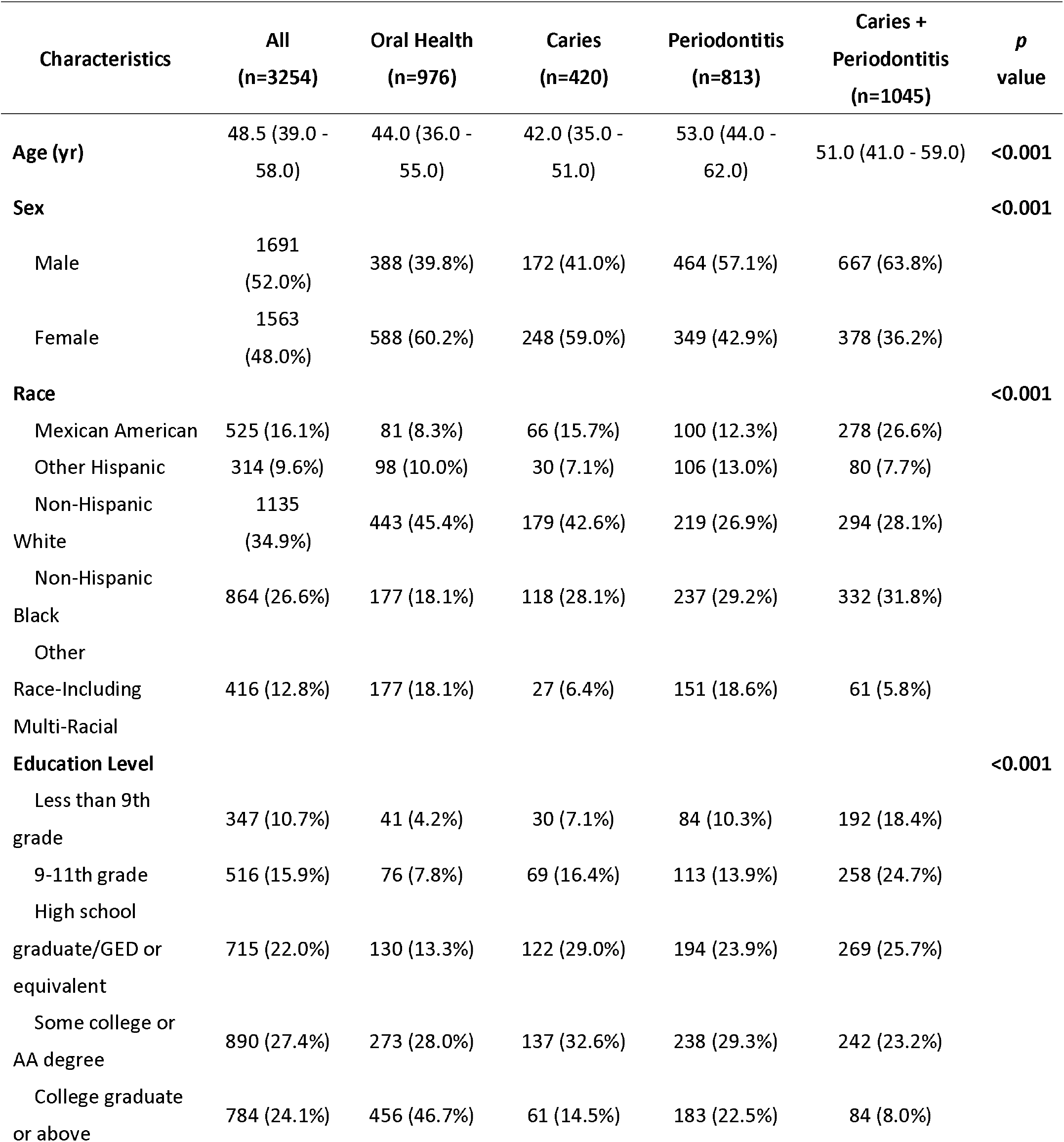
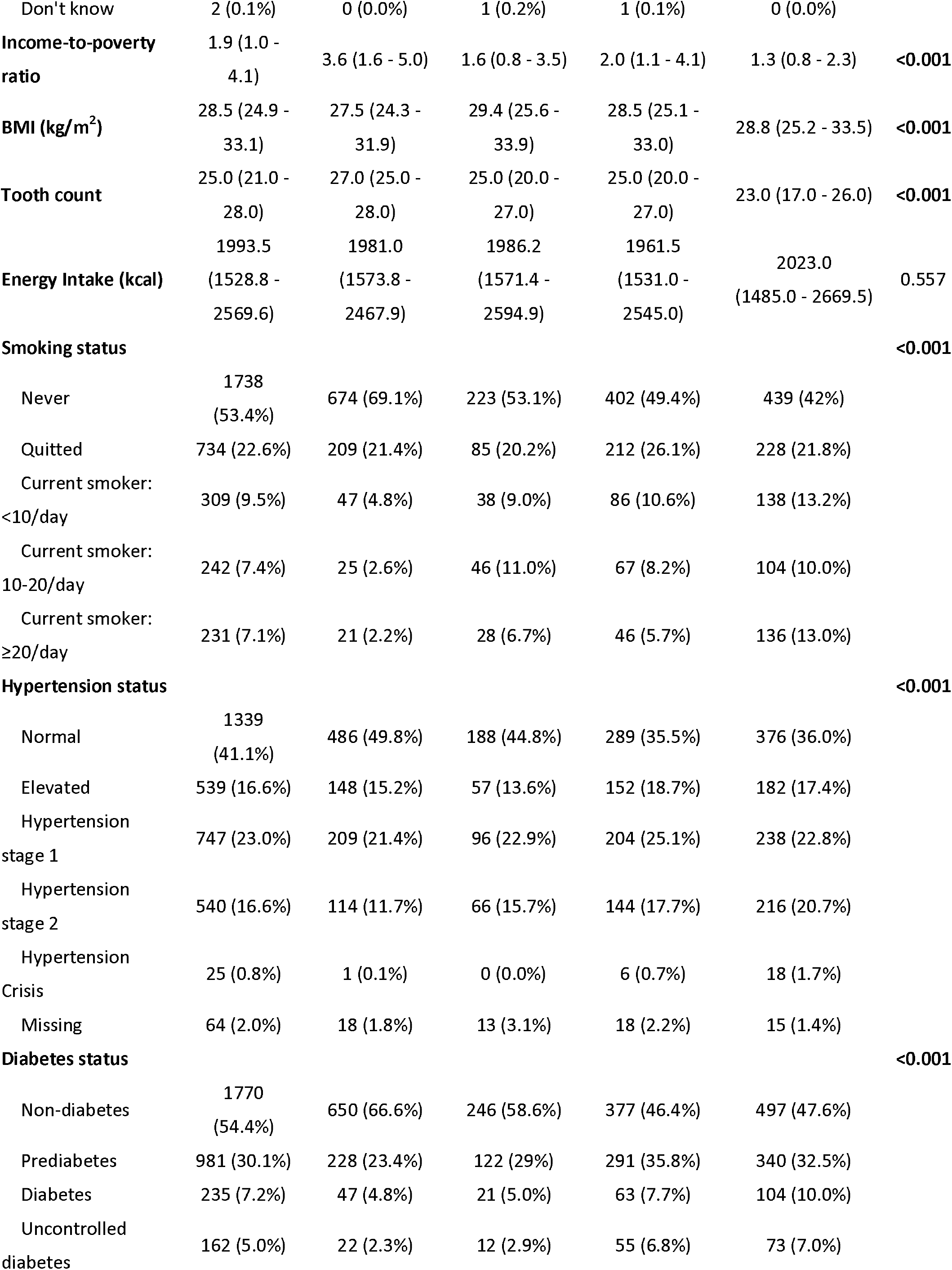
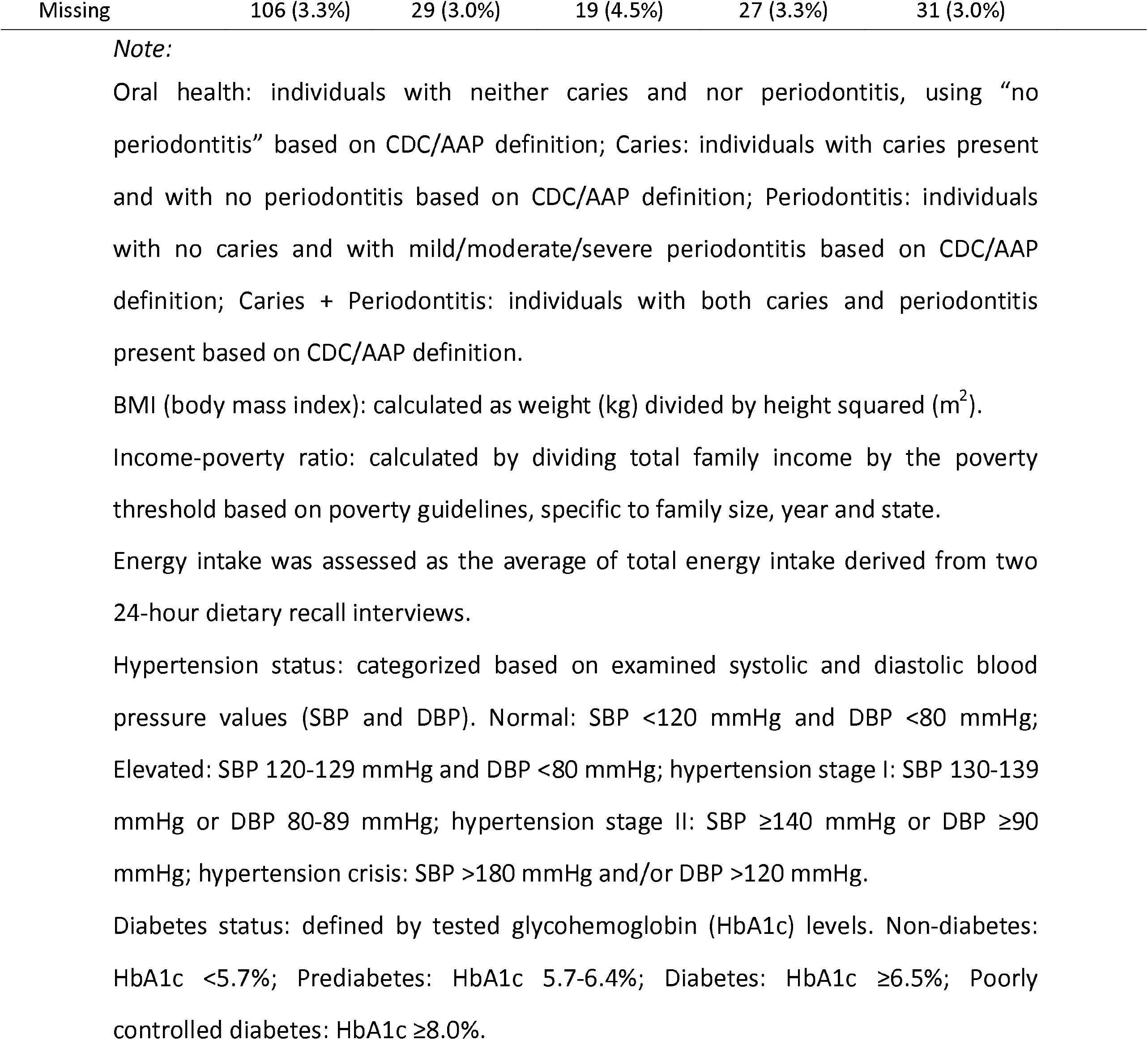
Characteristics of study participants with different oral health conditions based on caries and periodontitis status with CDC/AAP periodontitis definitions.

### Dietary Intake is Associated with Oral Rinse Microbial Community Structure Across Oral Conditions

Overall, dietary intake, considered as a combined set of variables, explained 3.6% of the variance in oral rinse microbial community structure (*p* < 0.001), comparable to oral disease status (R^2^=3.5%) or smoking (R^2^=2.8%), and greater than other sociodemographic factors **(Figure 1B)**.

Dietary indices were significantly associated with microbial community structure in the overall population and within individual oral condition groups **(Figure 1C** and **Appendix Figure 2)**. The Healthy Eating Index 2020 (HEI2020), the Dietary Approaches to Stop Hypertension (DASH) index, and the Mediterranean diet score (MED) showed directionally concordant associations with microbial community structure, whereas DII showed the opposite pattern. In stratified analyses, HEI2020 and MED explained the greatest microbial variance in the caries-only group (HEI2020: R^2^=0.31%, MED: R^2^=0.49%; both *p* < 0.01), whereas DASH and DII explained the greatest variance in the periodontitis-only group (DASH: R^2^=0.26%, DII: R^2^=0.31%; both *p* < 0.01).

Individual energy-adjusted food variables explained smaller proportions of microbial variance (R^2^ ≈ 0.1-0.5%) but showed distinct directional patterns **(Figure 1D)**. Added sugars, solid fats, potatoes, and alcoholic drinks associated with microbial community structure in a similar direction, whereas generally health-promoting foods (e.g. whole grains, nuts and seeds, vegetables, and fruits) showed the opposite direction. Seafood, meat, poultry, starchy vegetables, beans, and refined grains formed a third heterogeneous pattern.

After stratification by oral condition, fewer food variables remained significant (20 of 25 significant in at least one group), while the overall directional structure was largely preserved **(Figure 1E)**. Among oral health individuals, solid fats were aligned with the direction of association of fruits and vegetables. In the caries-only group, fewer dietary variables remained significant. In periodontitis-only and co-existing disease groups, added sugars, cheese, milk, and total dairy showed similar directions, whereas poultry, seafood, refined grains, and beans aligned more closely with fruits and vegetables.

### Distinct Diet-Microbiome Association Patterns Independent of Oral Conditions

Clustered heatmap analysis revealed structured diet-microbiome association patterns **(Figure 2A)**. Overall, vegetables and fruits, together with fresh animal-based foods and higher HEI2020, DASH, and MED scores, were positively associated with taxa commonly linked to oral health and inversely associated with taxa frequently linked to caries or periodontitis. Fresh animal products showed a weak significant correlation with total vegetable intake **(Appendix Figure 3)**. In contrast, added sugars, alcoholic drinks, cured meat, potatoes, dairy products, and higher DII scores, showed the opposite pattern. Additionally, microbial group 2, composed predominantly of periodontal health-associated taxa (e.g. *Kingella, Streptococcus, Rothia, Veillonella*), showed a partially distinct association profile. This group was negatively associated with plant-based carbohydrate-rich foods (beans, grains, starchy vegetables) but positively associated with cured meat, nuts and seeds, and dairy products.

**Figure 2.**
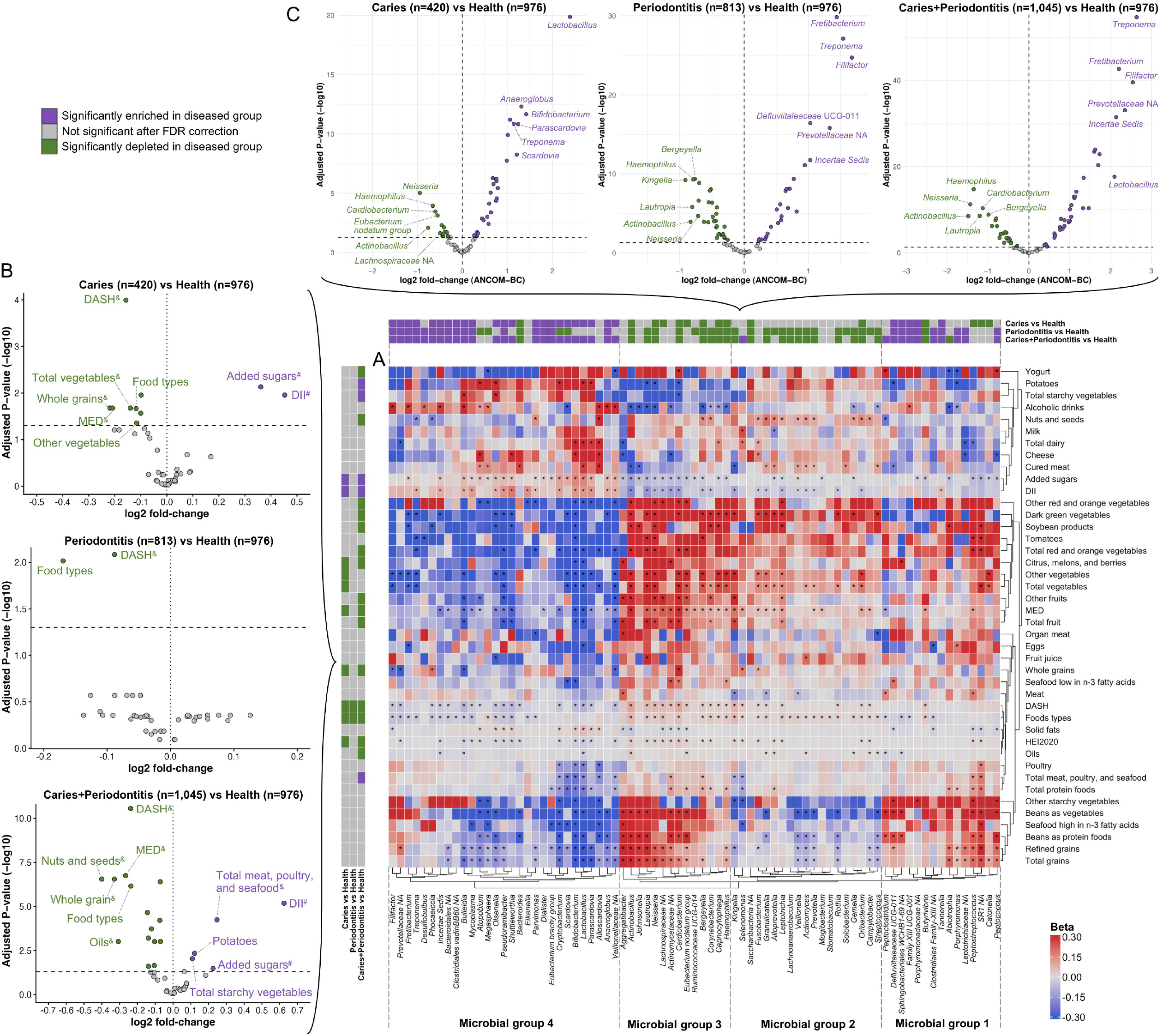
Associations between dietary intakes and oral rinse microbial taxa independent of oral conditions. (A) Main heatmap summarizing associations between dietary intakes and oral rinse microbial taxa abundance independent of oral conditions. Associations were estimated using MaAsLin2, adjusting for age, sex, and oral conditions. Beta coefficients are shown, and statistical significance after FDR correction is indicated by * when adjusted *p* < 0.05. Dietary variables and microbial taxa were hierarchically clustered here based on their MaAsLin2 association patterns, and distinct microbial groups were identified based on the clustered heatmap. (B) Volcano plots showing differences in dietary intake between each oral disease group (caries-only, periodontitis-only, or co-existing caries + periodontitis) and the oral health reference group. Group differences were evaluated using Mann-Whitney U tests on log1p-transformed intake values. Effect sizes are presented as log_2_-transformed mean differences between groups, and statistical significance was determined using Benjamini–Hochberg false discovery rate (FDR) correction. The horizontal dashed line indicates adjusted *p* = 0.05, and the vertical dashed line denotes no difference between groups. Each point represents one dietary variable, the green points indicate dietary variables significantly enriched in the oral health group, whereas purple points indicate dietary variables significantly enriched in the diseased group. Variables with the 6 largest absolute log_2_ fold-changes are labeled, with the superscript “&” indicates encouraged or health-promoting intake according to Food Patterns Equivalents Database (FPED)-based dietary recommendations, whereas superscript “#” indicates intake suggested to be limited or considered pro-inflammatory intake in FPED recommendation. The accompanying mini-heatmap highlights dietary variables with significant differences across group comparisons, and is aligned with the main heatmap. (C) Volcano plots showing differential abundance of oral rinse microbial taxa between each oral disease group and the oral health reference group. Differential abundance was assessed using ANCOM-BC on read count data. Effect sizes are presented as log_2_ fold changes between groups, and statistical significance was determined using FDR correction. The horizontal dashed line indicates adjusted *p* = 0.05, the vertical dashed line denotes no difference between groups. Each point represents one microbial taxon, the green points indicate taxa significantly enriched in the oral health group, whereas purple points indicate taxa significantly enriched in the diseased group. Taxa with the 6 largest absolute fold changes are labeled. The accompanying mini-heatmap highlights taxa with significant differential abundance across group comparisons, and is aligned with the main heatmap. For panels (B) and (C), the mini-heatmaps use grey indicates no significant difference, green indicates higher intake/abundance in the oral health group, and purple indicates higher intake/abundance in the diseased group.

To further contextualize these patterns, distribution differences in dietary intake **(Figure 2B)** and microbial taxa **(Figure 2C)** were compared between diseased groups and the oral health group.

Individuals with periodontitis only showed lower DASH scores and reduced dietary diversity, whereas caries-only and co-existing disease groups showed less favorable dietary profiles characterized by higher DII scores and added sugar intake, lower HEI2020, MED, and DASH scores, and lower intake of whole grains and fruits. The co-existing disease group additionally showed higher intake of starchy vegetables and animal-based foods, and lower intake of oils, fruits, nuts and seeds, and yogurt.

Differential abundance analysis identified taxonomic differences between diseased groups and oral health, broadly consistent with previous findings (Xie et al. 2026), characterized by enrichment of disease-associated taxa and depletion of health-associated taxa.

### Oral Rinse Microbial Groups Mediate Diet-Oral Disease Associations

Based on clustered diet-microbiome association profiles, oral rinse taxa were classified into four coordinated microbial groups **(Figure 2A)**, representing predominantly periodontitis-associated proteolytic anaerobes (microbial group 1), saccharolytic and/or biofilm-forming taxa (group 2), health-associated facultative anaerobic and aerobic commensals (group 3), and dysbiosis-associated acidogenic and inflammatory anaerobes (group 4). The full taxonomic composition of each microbial group is provided in **Appendix Table 1**.

Parallel mediation models, including all four microbial groups, were used to evaluate indirect associations between dietary intake exposure and oral condition outcomes. **Figure 3** showed representative models for total vegetable intake and added sugars. For the outcomes of caries versus oral health **(Figure 3A)** and co-existing disease versus health **(Figure 3C)**, microbial group 2 consistently demonstrated significant indirect effects in opposite directions for total vegetable and added sugar exposures. However, other microbial groups with significant indirect effects differed across oral disease outcomes. For vegetable intake, microbial groups 2 (*path b* = -0.280) and 3 (*path b* = -0.353) showed significant indirect effects for caries, whereas microbial groups 2 (*path b* = -0.271) and 4 (*path b* = 0.327) were significant for co-existing disease. For added sugar intake, microbial groups 1 (*path b* = -0.305), 2 (*path b* = -0.282), and 3 (*path b* = -0.352) showed significant indirect effects for caries, whereas microbial groups 2 (*path b* = -0.273) and 4 (*path b* = 0.330) were significant for co-existing disease. For periodontitis versus health **(Figure 3B)**, the total effect was not significant, so the mediation results were not further interpreted. Complete mediation results are provided in **Appendix Table 3-5**.

**Figure 3.**
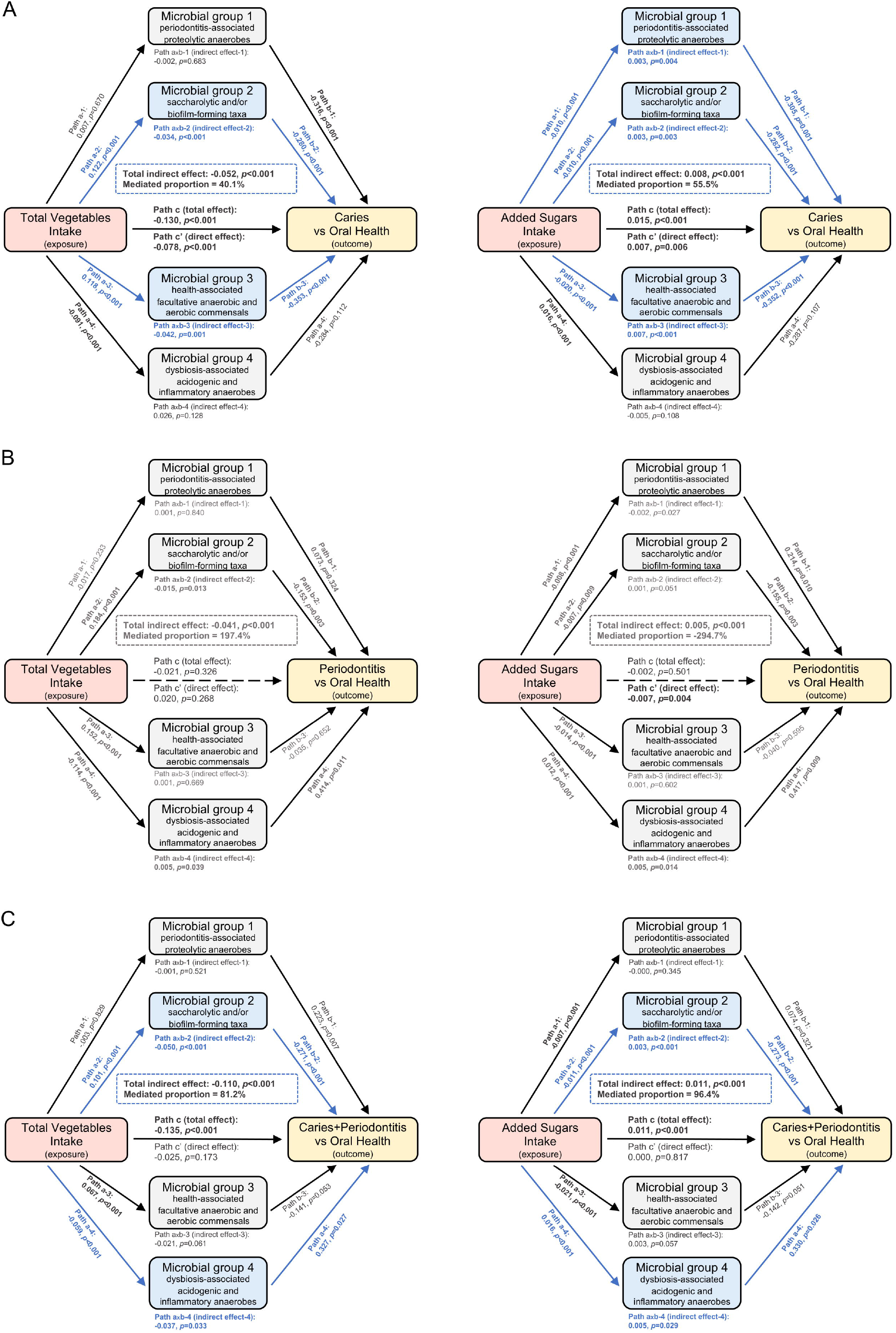
Multiple parallel mediation analyses linking dietary intake, oral rinse microbial groups, and oral conditions. (A) Multiple parallel mediation models evaluating whether oral rinse microbial groups mediate the associations between representative dietary exposures (total vegetable intakes and added sugars) and caries (vs. oral health). (B) Multiple parallel mediation models evaluating whether oral rinse microbial groups mediate the associations between representative dietary exposures and periodontitis (vs. oral health). (C) Multiple parallel mediation models evaluating whether oral rinse microbial groups mediate the associations between representative dietary exposures and co-existing caries + periodontitis (vs. oral health). For panels (A-C), dietary exposures were energy-adjusted. Microbial groups were defined based on the clustered heatmap shown in Figure 2. For each microbial group, the mediator represents the Z-score standardized mean CLR-transformed abundance of taxa within that group. The taxonomic composition of microbial group 1-4 is provided in Appendix Table 1. All models were specified as multiple parallel mediation models including four mediators simultaneously. *Path a* represents the association between the dietary exposure and each microbial group, and path *b* represents the association between each microbial group and the binary oral condition outcome conditional on the exposure. The *indirect effect* for each mediator was calculated as the product of *paths a* and *b*, and the *total indirect effect* equals the sum of indirect effects across all mediators. The *total effect* represents the overall association between the dietary exposure and the outcome, and the *direct effect* (clZ) represents the association after accounting for all mediators. For *path a*, positive estimates indicate that higher exposure levels are associated with higher microbial group scores (i.e., greater group abundance), whereas negative estimates indicate lower scores; *For path b*, positive estimates indicate that higher microbial group scores are associated with increased odds of the disease outcome relative to oral health, whereas negative estimates indicate decreased odds. For the *indirect, direct, and total effects*, positive estimates indicate that higher exposure levels are associated with increased odds of the disease outcome relative to oral health, whereas negative estimates indicate decreased odds. Results from additional mediation analyses involving dietary indices and other individual food intake exposures across different oral condition outcomes are presented in Appendix Table 3-5.

### Unsupervised Dietary Patterns Stratify Oral Microbial Profiles

K-means clustering of energy-adjusted food variables identified three population-level dietary clusters **(Figure 4A)**, with different dietary indices distribution **(Figure 4B)** and food intake profiles **(Figure 4C)**: an animal-based and pro-inflammatory pattern (cluster 1), a mixed grain-dairy pattern (cluster 2), and a plant-rich pattern (cluster 3). Among them, cluster 3 showed the highest HEI2020, DASH, and MED scores and the lowest DII values.

**Figure 4.**
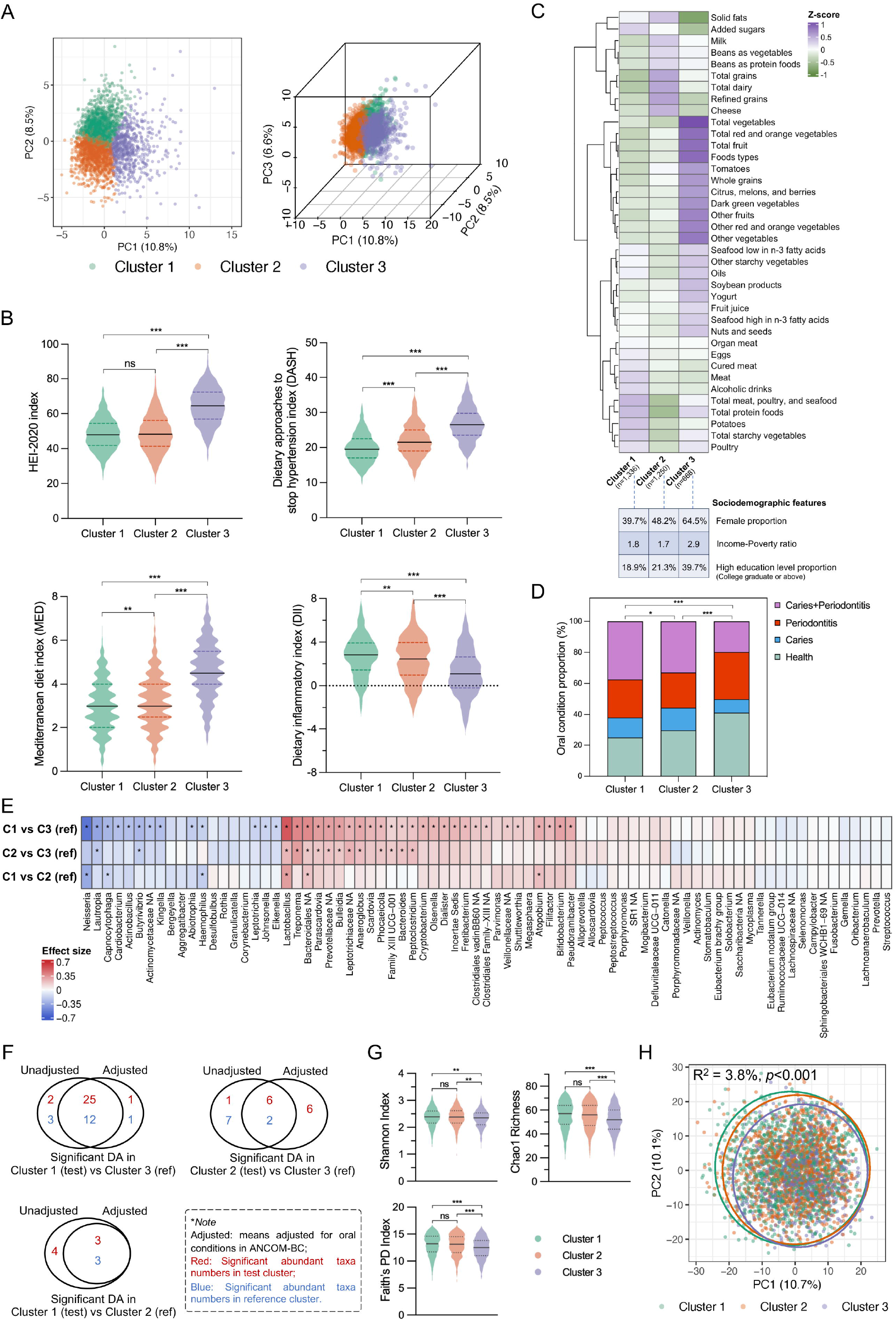
Dietary intake patterns defined by unsupervised machine learning explain oral rinse microbial differences independent of oral conditions. (A) Distribution of dietary intake clusters identified by unsupervised machine learning visualized in two-dimensional and three-dimensional principal component analysis (PCA) plots of food intake variables. (B) (B) Distribution of dietary indices across dietary intake clusters. Differences were assessed using the Kruskal-Wallis test, followed by Dunn’s test for post hoc pairwise comparisons. (C) Heatmap of energy-adjusted food intake patterns and representative sociodemographic characteristics across dietary intake clusters. Food intakes were energy-adjusted per 1,000 kcal and standardized as Z-scores relative to the overall population mean. Colors represent standardized differences (cluster mean minus overall mean divided by the standard deviation), indicating cluster-specific deviations from the population average. Representative sociodemographic characteristics including the proportion or median value of sex, income and education level for each dietary cluster, which are shown below the heatmap. Full population characteristics of the dietary clusters are presented in Appendix Table 2. (D) Distribution of oral conditions across dietary intake clusters. Differences were evaluated using the Chi-square test. (E) Heatmap of differentially abundant microbial taxa across dietary intake cluster comparisons. Differential abundance was evaluated using ANCOM-BC adjusted for oral conditions. Log_2_ fold-change effect sizes were visualized, and statistical significance after FDR correction is denoted as * (adjusted *p* < 0.05). C1, C2, and C3 represent dietary intake clusters 1, 2, and 3, respectively. In comparisons C1 vs C3 and C2 vs C3, cluster 3 was used as the reference; in C1 vs C2, cluster 2 was used as the reference. (F) Venn diagrams summarizing overlap of differentially abundant taxa identified by unadjusted ANCOM-BC and ANCOM-BC adjusted for oral conditions across dietary intake cluster comparisons. Red numbers indicate taxa abundant in the test cluster, and blue numbers indicate taxa abundant in the reference cluster. (G) Distribution of microbial alpha diversities across dietary intake clusters. Differences were assessed using the Kruskal-Wallis test, followed by Dunn’s test for post hoc pairwise comparisons. (H) PCA of oral rinse microbial community structure based on Aitchison distance across dietary intake clusters. Each point represents one sample; ellipses indicate 95% confidence regions. Explained variance (R^2^) and *p* values were obtained using sequential permutational multivariate analysis of variance (PERMANOVA). A sequential model was applied in which oral conditions were entered first to account for disease-associated microbial variation; the R^2^ for dietary intake clusters represents the variance explained after partitioning the effect of oral conditions. For panel (B), (D), and (G), statistical significance is denoted as ns: no significance, *: *p* <0.05, **: *p* <0.01, ***: *p* <0.001.

In addition, these dietary clusters differed in sociodemographic characteristics and in their oral health status **(Figure 4C-D)**. Cluster 3 included a higher proportion of females (64.5%), higher educational attainment (39.7% college graduates or higher), and higher income (median income-poverty ratio of 2.9). Detailed population characteristics across dietary clusters are provided in **Appendix Table 2**. Across clusters, the proportion of oral health increased from cluster 1 to cluster 3, whereas the proportion of co-existing disease decreased. The proportion of caries with or without periodontitis was lowest in cluster 3, whereas the proportions of health and periodontitis-only were highest.

Microbial profiles across dietary clusters were evaluated with adjustment for oral conditions **(Figure 4E)**. Compared with cluster 3, cluster 1 showed lower abundance of nitrate-reducing commensals (e.g. *Neisseria, Haemophilus, Capnocytophaga, Butyrivibrio, and Leptotrichia*) and higher abundance of disease-associated acidogenic and proteolytic taxa (e.g. *Scardovia, Parascardovia, Lactobacillus, Bifidobacterium, Filifactor, Fretibacterium, and Treponema*). Fewer taxa differed between clusters 2 and 3, while cluster 2 similarly showed lower abundance of nitrate-reducing taxa and higher abundance of disease-associated taxa. Relative to cluster 2, cluster 1 showed a more pronounced anaerobic and acidogenic profile, with lower abundances of *Neisseria, Haemophilus*, and *Capnocytophaga*, and higher abundances of *Lactobacillus, Atopobium*, and *Bacteroidales* NA.

These differential abundance patterns were largely consistent with and without adjustment for oral conditions, supporting the robustness of microbial differences across dietary patterns **(Figure 4F)**. Microbial alpha diversity did not differ between clusters 1 and 2, whereas cluster 3 showed significantly lower Shannon, Chao1, and Faith’s PD **(Figure 4G)**. Sequential PERMANOVA **(Figure 4H)** showed that dietary clusters explained an additional 3.8% of beta diversity variance after accounting for oral conditions (*p* < 0.001).

## Discussion

Diet is considered a key modulator of the human microbiome, particularly in the gut, where most nutrients are metabolized (Zmora et al. 2019). As the initial site of dietary exposure, the oral cavity can also be influenced by diet, yet existing studies are often based on relatively small samples and focused on limited components, particularly carbohydrates or vegetables. In this population-level study, a broader spectrum of food components was associated with oral rinse microbiota composition. Notably, the collective contribution of these food components explained a proportion of variance (3.6%) in the oral rinse microbial community comparable to oral disease status or smoking, highlighting diet as an important ecological factor in the oral environment.

Since both dietary intake and oral microbiome composition are strongly associated with oral diseases (Shen et al. 2023; Xie et al. 2026), oral conditions should be considered when evaluating diet-microbiome relationships. Oral conditions were therefore addressed through stratification and statistical adjustment, or were treated as hypothetical outcomes in mediation models.

In this study, diet-microbiome associations involving carbohydrates and vegetables were consistent with previous findings. Added sugar and green vegetables showed opposite association patterns with the oral rinse microbiota at both community and taxon levels. For example, *Rothia* and *Neisseria* were positively associated with vegetable intake, whereas *Lactobacillus* and *Bifidobacterium*, were positively associated with added sugar intake and enriched in caries individuals. These findings align with existing evidence showing that frequent intake of fermentable carbohydrates favors acidogenic and acid-tolerant taxa and promotes acidic microenvironments linked to dental caries (Marsh 2003; Pitts et al. 2017). In contrast, vegetables may support oral health through alternative metabolic pathways (Wright et al. 2020), particularly via nitrate metabolism (Rosier et al. 2022), which may limit oral acidification (Rosier et al. 2021) and generate nitric oxide with antimicrobial and anti-inflammatory properties(Rosier et al. 2025). These concordant patterns support the biological plausibility of the observed associations and lend credibility to the analytical approach.

Interestingly, fresh animal-based foods (red meat, poultry, and seafood) showed microbial association patterns more similar to those of vegetables and fruits than expected. Although the metabolization of host proteins by subgingival communities is associated with the dysbiosis found in periodontal diseases (Takahashi 2015) and red meat has been linked to systemic inflammatory markers related to periodontitis (DeMayo et al. 2021). The observed associations could suggest that fresh animal products are part of healthier diets in this population and/or that pro-inflammatory effects of animal-based foods may act primarily through systemic pathways without negatively effecting the oral rinse microbiota (Wang et al. 2025). Cured meats, on the other hand, showed distinct associations compared with fresh animal-based foods and were more positively linked to disease-associated bacteria/oral disease.

Dairy products, particularly milk and cheese, showed association patterns similar to those of cured meat and added sugar. Although dairy intake is not strongly associated with systemic inflammation (Hess et al. 2021), lactose and other fermentable substrates may support saccharolytic taxa. Notably, dietary correlation analysis did not show a strong positive correlation between dairy and added sugar consumption **(Appendix Figure 3)**, suggesting that these associations are unlikely to be explained by co-intake patterns.

Carbohydrate-containing plant foods such as beans, grains, and non-potato starchy vegetables showed heterogeneous association patterns compared with added sugar or green and red vegetables. These foods contain resistant starch and complex polysaccharides, which differ in fermentability and metabolic accessibility from simple sugars (Slavin 2013). While potatoes showed partially distinct patterns relative to other starchy vegetables, these possibly relate to their relatively high glycemic index and frequent consumption in processed forms (Muraki et al. 2016). These findings suggest that plant-derived carbohydrate sources may differ in their associations with oral bacteria depending on carbohydrate structure and degree of processing.

Dietary indices such as HEI2020, DASH, MED, and DII summarize overall diet quality or inflammatory potential and have been associated with oral microbial composition (Augimeri et al. 2024; Yue et al. 2025b). However, predefined indices may not fully capture real-world dietary behaviors. Using unsupervised clustering, three major dietary patterns were identified: an animal-based and pro-inflammatory pattern (cluster 1), a mixed grain-dairy pattern (cluster 2), and a plant-rich pattern (cluster 3). These patterns were associated with distinct sociodemographic characteristics, suggesting that dietary behaviors reflect broader lifestyle and social contexts (Stein et al. 2021). Individuals following the plant-rich pattern showed a higher proportion of oral health and microbial profiles enriched with nitrate-reducing taxa (e.g. *Neisseria, Haemophilus*). In contrast, dietary patterns characterized by higher intake of animal-based or processed foods were more frequently associated with proteolytic or acidogenic taxa commonly linked to dysbiosis. These findings support the view that diet-microbiome relationships reflect combined effects of multiple dietary exposures rather than isolated nutrients.

Diet-microbiome associations also varied across oral disease conditions. In mediation analyses, microbial groups showing significant indirect effects differed across disease outcomes, suggesting that host-microbiome responses to dietary exposures may vary across disease contexts or host genetic backgrounds (Kamitaki et al. 2026). In our study, the plant-rich pattern showed the lowest caries and caries + periodontitis proportion but a relatively higher periodontitis proportion. This contrast may reflect differences in disease mechanisms: caries is strongly influenced by local effects of carbohydrate fermentation by dental biofilms (without fermentable carbohydrates, there would not be dental caries), whereas periodontitis may have a pathogenesis, which is more strongly associated with systemic inflammation and genetic susceptibility (Schaefer et al. 2025). Further studies are needed to clarify these relationships.

Several limitations should be considered. First, the cross-sectional design precludes causal inference; longitudinal studies are needed to evaluate whether dietary exposures directly influence microbial dynamics and oral disease development. Second, although oral rinse samples represent composite oral microbial communities, they may preferentially reflect tongue-associated taxa (e.g. *S. salivarius and Rothia mucilaginosa*)(Wilbert et al. 2020) and may not fully capture site-specific supragingival or subgingival niches, limiting interpretation of localized mechanisms. Third, the lack of access to raw sequencing data (e.g., FASTQ files) in NHANES and the availability of microbiome data only at the genus level limit species-level resolution and restrict associations between bacterial taxa and other parameters. Finally, dietary intake was assessed using 24-hour dietary recalls and may be subject to reporting bias.

This study extends previous work by examining diet-microbiome associations in a large nationally representative population and by integrating dietary indices, individual food components, population-level dietary patterns, microbial community structure, and oral disease outcomes within a unified analytical framework.

Overall, dietary intake was associated with oral microbiota composition and oral health conditions in this nationally representative population. Distinct diet-microbiome association patterns and microbial mediation signals suggest links among dietary behaviors, oral bacterial profiles, and oral disease risk. Longitudinal and mechanistic studies are needed to clarify the direction and causality of these relationships.

## Data Availability

All data produced in the present study are available upon reasonable request to the authors

## Author contribution

Y. Xie, contributed to conception, design, data acquisition and interpretation, drafted and critically revised the manuscript; M. Bi, contributed to data acquisition, analysis, and critically revised the manuscript; W. Gu, Y. Li, contributed to data analysis, and critically revised the manuscript; A. Roccuzzo, contributed to data interpretation, and critically revised the manuscript; B.T. Rosier, contributed to conception, design, data interpretation, and critically revised the manuscript; and M.S. Tonetti, contributed to conception, design, data analysis and interpretation, drafted and critically revised the manuscript. All authors gave their final approval and agree to be accountable for all aspects of the work.

## Data availability

The data used in this study are publicly available from the National Health and Nutrition Examination Survey (NHANES) database and can be accessed at https://wwwn.cdc.gov/nchs/nhanes/. The code used for the statistical analyses is available at https://github.com/YuX-97/NHANES-diet-microbiome.

