## Appendix for "Diet Explains Significant Variance in Oral Microbial Community Structure"

**Appendix Methods**

**Study population**

The United States National Health and Nutrition Examination Survey (NHANES) database was used in this study. NHANES is a nationally representative, cross-sectional survey conducted in two-year cycles to assess the health and nutritional status of the non-institutionalized U.S. civilian population, using a complex, multistage probability sampling design. Data collection includes standardized in-home interviews and clinical examinations performed in Mobile Examination Centers (MECs). The NHANES protocol was approved by the National Center for Health Statistics Ethics Review Board, and detailed survey procedures are publicly available (https://www.cdc.gov/nchs/nhanes/index.htm).

This study utilized data from the 2009-2010 and 2011-2012 NHANES cycles, which included oral rinse samples with 16S rRNA gene sequencing data, full-mouth clinical examinations, and two 24-hour dietary recalls. Participants were excluded if they were: (i) younger than 18 years; (ii) pregnant; (iii) lacking complete full-mouth dentition examination data; (iv) having fewer than two natural teeth; or (v) having incomplete or unreliable dietary recall data. The participant selection process is illustrated in **Appendix Figure 1**.

**Clinical examination**

Oral health examinations were conducted by licensed dentists in the MECs. Examinations were directly recorded by trained health technicians using a computerized data collection system. All examinations were performed in a dedicated MEC dental room equipped with a portable dental chair, dental light, and compressed air, following standardized NHANES protocols(Centers for Disease Control and Prevention and Statistics 2024a; 2024b).

Participants first underwent a tooth-count assessment, followed by caries evaluation. For this study, individual was classified as having dental caries if at least one decayed tooth was recorded in the NHANES database. Caries assessment protocols differed slightly between cycles. In the 2009-2010 cycle, only a binary indicator for the presence of at least one decayed tooth was available, and detailed examination procedures were not documented in the public manual. In the 2011-2012 cycle, a standardized caries examination protocol was described in the NHANES Oral Health Examiners Manual. All teeth except third molars were assessed. Quadrants were dried with air as needed, and examiners used a surface-reflecting mirror and a No. 23 explorer. Frank lesions were defined as gross cavitation. For anterior teeth, lingual, facial (buccal), mesial, and distal surfaces were examined; for posterior teeth, lingual, occlusal, facial (buccal), mesial, and distal surfaces were evaluated. Incipient proximal lesions required detection of an enamel surface break with the explorer in posterior teeth, and either explorer-detected enamel break or visualization by transillumination in anterior teeth.

Periodontal examination protocols were consistent across both cycles. Examinations were conducted by the dental examiners for full-mouth excluding third molars, six sites per tooth (distofacial, midfacial, mesiofacial, distolingual, midlingual, and mesiolingual) were assessed with a periodontal probe. Specifically, Probing pocket depth and gingival recession were measured during examination, while clinical attachment loss was calculated as probing pocket depth minus gingival recession.

Oral condition categories were further defined based on caries and periodontitis status and classified into four groups: (i) oral health (neither caries nor periodontitis present), (ii) caries (only dental caries present, without periodontitis), (iii) periodontitis (only periodontitis present, without dental caries), and (iv) co-existing caries and periodontitis (both caries and periodontitis present). Periodontitis was defined using the Centers for Disease Control and Prevention–American Academy of Periodontology (CDC/AAP) classification(Eke et al. 2012). The Application of the 2018 Periodontal Status Classification to Epidemiological Survey data (ACES) system (Holtfreter et al. 2024) was not used, as no participants were classified as “no periodontitis” under that framework in the analyzed cycles.

**Dietary intake assessment**

Dietary intake variables included dietary indices, Food Patterns Equivalents Database (FPED) food pattern components, consumed food types, and total energy intake. Dietary intake data were derived from two non-consecutive 24-hour dietary recalls, and the mean intake across the two recalls was used for further analysis.

Dietary indices were calculated using the dietaryindex R package (Zhan et al. 2024), applying published scoring algorithms for the Healthy Eating Index 2020 (HEI2020) (Kemp et al. 2022; Kennedy et al. 1995), the Dietary Approaches to Stop Hypertension (DASH) index (Appel et al. 1997), the alternate Mediterranean diet (aMED) score (Trichopoulou et al. 2003), and the Dietary Inflammatory Index (DII) (Shivappa et al. 2014).

HEI2020 assesses adherence to the 2020-2025 Dietary Guidelines for Americans, with higher scores indicating better adherence. The DASH index reflects adherence to the Dietary Approaches to Stop Hypertension pattern which is rich in fruits, vegetables, whole grains, and low-fat dairy, and low in sodium and red meat. The aMED score evaluates adherence to a Mediterranean-style dietary pattern adapted for U.S. populations. For the HEI2020, DASH, and aMED index, higher scores indicate better adherence to healthy dietary patterns. The DII quantifies the inflammatory potential of the diet by standardizing intake values to a reference distribution, multiplying by literature-derived inflammatory effect scores, and summing across components; negative scores indicate a more anti-inflammatory diet, whereas positive scores reflect a more pro-inflammatory dietary profile.

For food pattern intake, reported individual foods in the 24-hour dietary recall were converted into 37 standardized food groups using FPED. The nine major FPED components include fruits, vegetables, and dairy (cup-equivalents); grains and protein foods (ounce-equivalents); added sugars (teaspoon-equivalents); solid fats and oils (gram-equivalents); and alcoholic drinks (number of drinks).

The consumed food types and total energy intake were directly obtained from the NHANES dietary datasets.

**Oral rinse collection and 16S rRNA Gene Sequencing**

Oral rinse samples were collected in the MECs prior to the oral health examination. Participants were instructed to rinse and gargle with 10 mL of saline mouthwash for 30 seconds. Samples were stored at −80°C until DNA extraction and sequencing, which were performed in certified laboratories according to NHANES protocols. Sequencing and primary data processing were centrally conducted by the NHANES program.

The V4 region of the 16S ribosomal RNA (rRNA) gene was amplified using primers 515F (5′-GTG CCA GCM GCC GCG GTA A-3′) and 806R (5′-GGA CTA CHV GGG TWT CTA AT-3′). Sequencing was performed on the Illumina HiSeq 2500 platform using 2 × 125 bp paired-end reads.

Raw reads were demultiplexed using QIIME 1 (version 1.9.1), generating separate forward and reverse FASTQ files for each participant. Due to insufficient overlap between paired-end reads, only forward reads were retained for downstream processing. Sequence processing was performed using the DADA2 pipeline (version 1.2.1), including quality filtering, error correction, chimera removal, and inference of amplicon sequence variants (ASVs). Taxonomic assignment was conducted using the SILVA v123 database without rarefaction. The final outputs included collapsed read count and relative abundance tables up to the genus level. As only collapsed read count and relative abundance tables were available in the public NHANES release, these datasets were used for the present analyses.

**Statistical analysis**

All statistical tests were two-sided with a significance level of α = 0.05. For analyses involving multiple comparisons, *p* values were adjusted using the Benjamini-Hochberg false discovery rate (FDR) procedure. All analyses were conducted in R version 4.4.1 (http://www.R-project.org, The R Foundation, Vienna, Austria).

Although NHANES recommends incorporating sampling weights for nationally representative inference, many microbiome analytical frameworks (e.g., compositional transformations, ANCOM-BC, MaAsLin2, dbRDA) are not designed to incorporate complex survey weights. Therefore, descriptive and microbiome analyses were conducted at the sample level without applying NHANES survey weights, to maintain internal methodological consistency across analyses.

For descriptive analyses of population characteristics and variable distributions across groups, normality of continuous variables was assessed using distributional diagnostics. As most continuous variables were non-normally distributed, they are presented as median and interquartile range (IQR), while categorical variables are presented as counts (n) and percentages (%). Group differences were evaluated using the Kruskal-Wallis test for continuous variables, followed by Dunn’s test with multiple-comparison correction for post hoc pairwise comparisons, and Pearson’s Chi-square test for categorical variables. Analyses were restricted to participants with complete data for variables included in each model.

- Microbiome Data Preprocessing

For oral rinse microbiome data, taxa were filtered to improve robustness and reduce sparsity. Taxa were retained if they had >10 reads in at least 30% of participants within at least one oral condition group (oral health, caries, periodontitis, or co-existing caries and periodontitis), and had taxonomic annotation at least to the Class level. Samples with fewer than 5,000 total reads were excluded. All downstream microbiome analyses were conducted using the filtered dataset.

- Dietary Intake Data Processing

FPED-derived food pattern intakes were energy-adjusted and expressed per 1,000 kcal to evaluate structural dietary composition independent of total energy intake. Dietary indices were analyzed according to their established scoring frameworks.

- Downstream Analysis

Given the compositional nature of sequencing data, compositional data analysis principles were applied. Read count tables were transformed using the centered log-ratio (CLR) transformation after appropriate zero handling.

For beta diversity analysis, Aitchison distances on centered log-ratio (CLR)-transformed data were calculated to assess microbial community dissimilarity. Permutational multivariate analysis of variance (PERMANOVA) was used to test statistical differences in overall microbial community structure across dietary intake, sociodemographic factors, and oral and general health conditions, with the coefficient of determination (R²) reported as the proportion of variance explained. Distance-based redundancy analysis (dbRDA) based on Aitchison distance was performed to evaluate the microbial community variance explained by dietary intake variables and to visualize the direction and magnitude of associations.

Differential abundance of microbial taxa between oral disease groups (caries, periodontitis, or co-existing disease) and the oral health reference group was assessed using Analysis of Compositions of Microbiomes with Bias Correction (ANCOM-BC), which accounts for compositionality and sampling bias.

Differences in dietary intake between each oral disease group and the oral health reference group were evaluated using Mann-Whitney U tests, and effect sizes for dietary intake comparisons were calculated as the log₂-transformed mean difference between groups.

Associations between dietary intake variables and microbial taxa were assessed using Microbiome Multivariable Association with Linear Models (MaAsLin2). CLR-transformed microbial abundance data were used as dependent variables. Models were adjusted for age, sex, and oral condition to attenuate potential confounding by disease status. FDR correction was applied within each model. Clustered heatmaps were used to summarize diet-microbiome association patterns. Based on the clusters of association patterns, microbial taxa were grouped into four microbial clusters.

To investigate whether microbial clusters mediated the association between dietary intake and oral disease, multiple parallel mediation models were constructed. Dietary indices or food pattern variables were defined as exposures. Binary oral disease status (each disease group versus oral health reference) was defined as the outcome. The four microbial cluster scores were included simultaneously as parallel mediators. Cluster scores were calculated as the z-score standardized mean CLR-transformed abundance of taxa within each cluster. The following effects were measured: (i) association between dietary intake and microbial cluster abundance (path a); (ii) association between microbial cluster abundance and oral disease outcome (path b); (iii) total effect of dietary intake on oral disease outcome (path c); (iv) separate and total indirect effects mediated through microbial clusters (separate and total path axb); and (v) direct effect of dietary intake on oral disease outcome independent of the microbiome (path c’). The proportion mediated was calculated as: β_indirect effect_ ⁄ β_total effect_ × 100.

To identify underlying dietary intake patterns within the study population, unsupervised machine learning approaches were applied. Energy-adjusted food pattern variables were first subjected to principal component analysis (PCA) for dimensionality reduction. Principal components with eigenvalues >1 were retained as input for clustering. Clustering tendency was evaluated using the Hopkins statistic (>0.8 indicated strong clustering structure). The optimal number of clusters was determined by integrating multiple metrics, including Jaccard index (>0.8 for cluster stability), multistart adjusted Rand index (ARI), and Silhouette index. Based on these criteria, k = 3 was selected for k-means clustering.

Cluster separation was visualized using PCA plots. Differences in dietary indices across clusters were assessed using the Kruskal-Wallis test with Dunn’s post hoc comparisons. Clustered heatmaps were used to visualize single food intake patterns across dietary clusters, with food intakes expressed per 1,000 kcal and standardized as Z-scores relative to the overall population mean. In addition, distribution of oral conditions across dietary clusters was compared using Chi-square tests.

Differential abundance of microbial taxa across dietary intake clusters was evaluated using ANCOM-BC, with and without adjustment for oral conditions. Overlap of differentially abundant taxa between adjusted and unadjusted models was visualized using Venn diagrams. Alpha diversity of Shannon index, Chao1 richness, and Faith's Phylogenetic Diversity (Faith’s PD) index across dietary intake clusters were compared using the Kruskal-Wallis test with Dunn’s post hoc comparisons. Beta diversity differences across dietary intake clusters were assessed using Aitchison distance and visualized through PCA. Statistical significance was evaluated using sequential PERMANOVA, with oral condition entered first in the model to account for disease-associated microbial variation. The R² attributed to dietary intake clusters therefore represents the proportion of variance explained after partitioning the effect of oral condition.

**Appendix Figures**

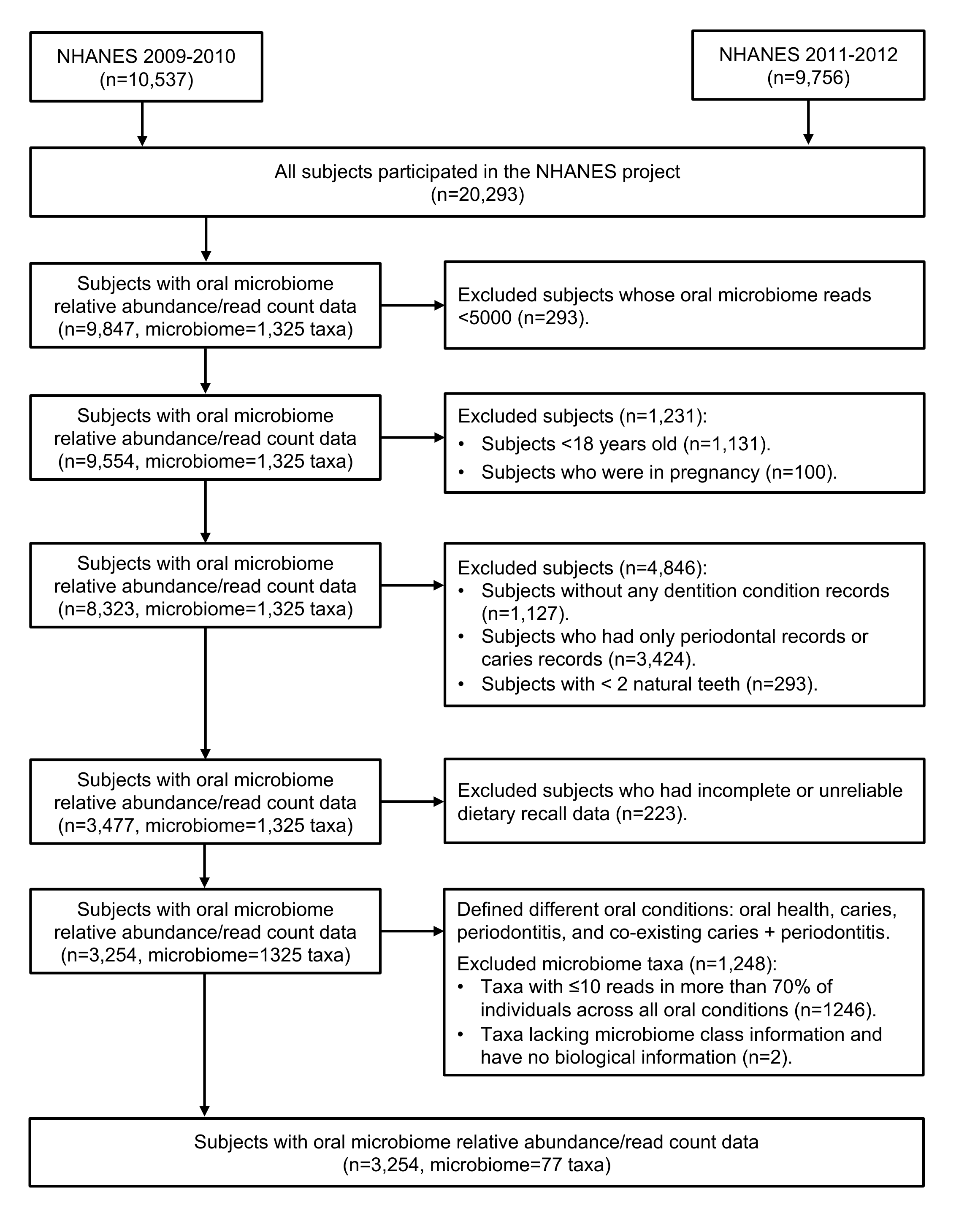

**Appendix Figure 1. Flow chart of subject and microbiome inclusion process from the NHANES 2009-2010 and 2011-2012 database.**

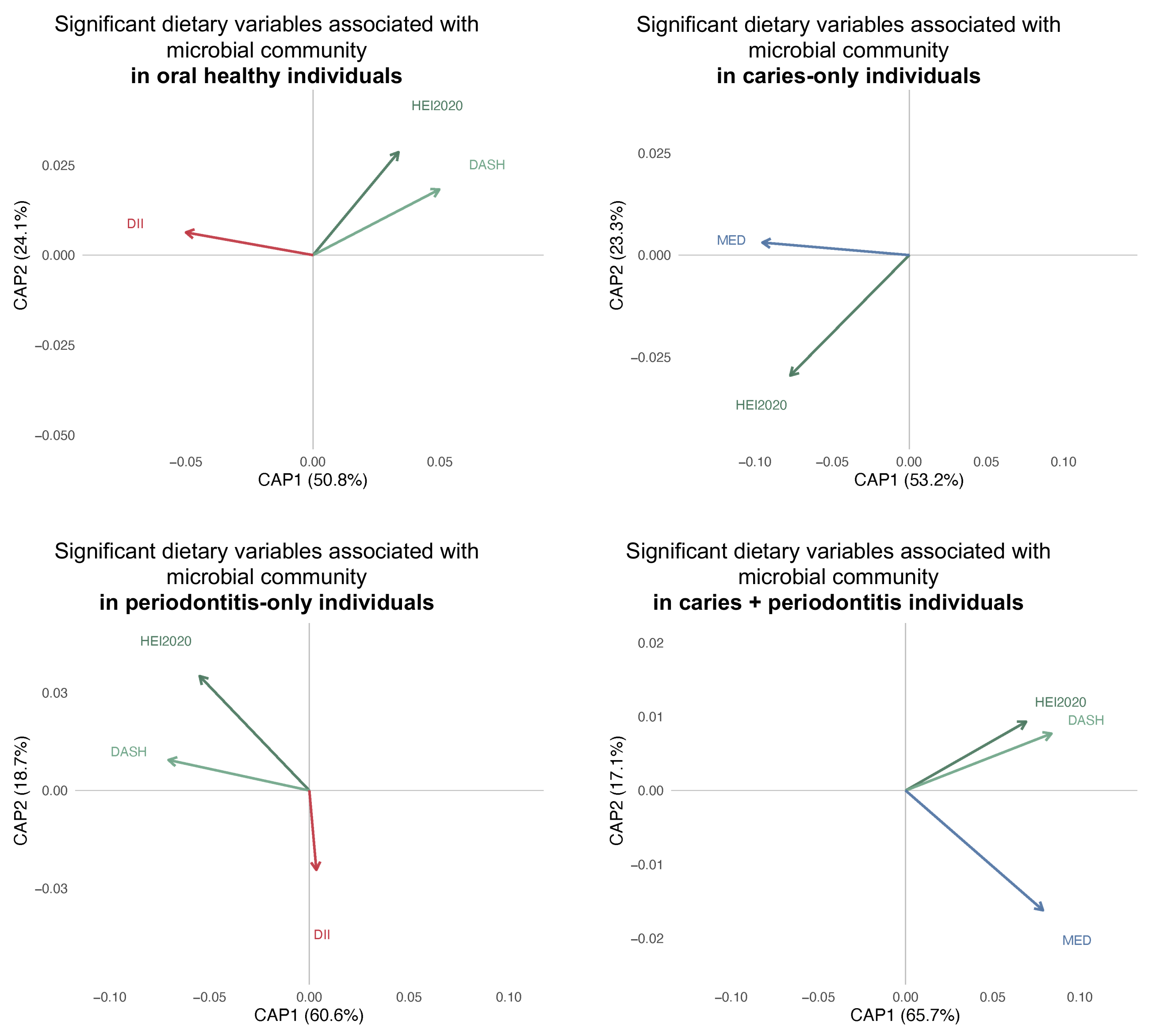

**Appendix Figure 2. Dietary indices associated with oral rinse microbial community structure across oral condition groups.**

Distance-based redundancy analysis (dbRDA) under Aitchison distance showing dietary indices significantly associated with variation in the overall oral rinse microbial community within each oral condition group. Arrows represent dietary indices significantly associated with microbial community structure. Arrow length reflects the strength of association with the constrained ordination axes. The absolute direction of an arrow is arbitrary; however, the relative angles between arrows indicate whether variables are associated with the microbial community in a similar or contrasting direction in the constrained ordination space. Variables with relative angles < 90° show associations in a similar direction, whereas variables with relative angles > 90° indicate associations in opposing directions.

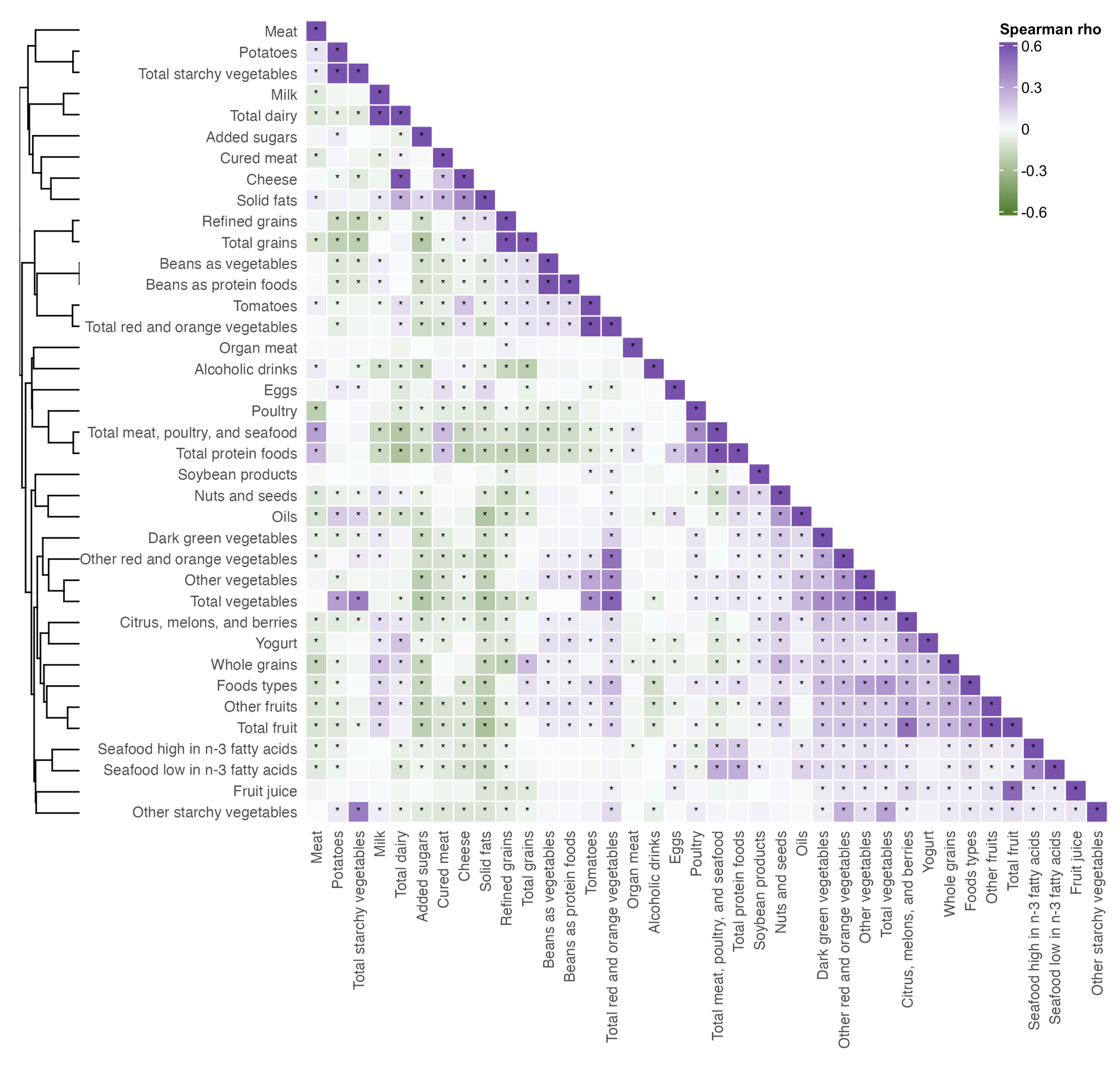

**Appendix Figure 3. Correlation structure among energy-adjusted food intake variables in the overall population.**

Pairwise correlations among food intake variables were evaluated using Spearman rank correlation coefficients based on energy-adjusted intake values (expressed per 1000 kcal). Positive correlations are shown in purple and negative correlations in green, with color intensity corresponding to the magnitude of the correlation coefficient. Statistical significance after false discovery rate (FDR) correction for multiple comparisons is denoted as * (adjusted *p* < 0.05).

**Appendix Tables**

**Appendix Table 1. Taxonomic composition of microbial clusters derived based on clustered heatmap in diet-microbiome MaAsLin2 association analysis.**

|  | **Taxa names** |
| --- | --- |
| **Microbial group 1**  **(proteolytic, anaerobic taxa associated with periodontitis)** | *Abiotrophia, Butyrivibrio, Family XIII UCG-001, Clostridiales Family-XIII NA, Peptoclostridium, Leptotrichiaceae NA, Porphyromonadaceae NA, Porphyromonas, Tannerella, Sphingobacteriales WCHB1-69 NA, SR1 NA, Defluviitaleaceae UCG-011, Catonella, Peptococcus, Peptostreptococcus* |
| **Microbial group 2**  **(saccharolytic and/or biofilm-forming taxa)** | *Rothia, Granulicatella, Leptotrichia, Kingella, Actinomyces, Alloprevotella, Prevotella, Gemella, Streptococcus, Mogibacterium, Lachnoanaerobaculum, Oribacterium, Stomatobaculum, Solobacterium, Selenomonas, Veillonella, Fusobacterium, Campylobacter, Saccharibacteria NA* |
| **Microbial group 3**  **(health-associated facultative anaerobic and aerobic commensals)** | *Actinomycetaceae NA, Corynebacterium, Bergeyella, Capnocytophaga, Johnsonella, Lautropia, Neisseria, Cardiobacterium, Actinobacillus, Aggregatibacter, Haemophilus, Eubacterium nodatum group, Lachnospiraceae NA, Ruminococcaceae UCG-014* |
| **Microbial group 4**  **(caries- and periodontitis-associated acidogenic and inflammatory anaerobes)** | *Eikenella, Desulfobulbus, Parascardovia, Bacteroidales NA, Prevotellaceae NA, Lactobacillus, Bulleidia, Treponema, Anaeroglobus, Veillonellaceae NA, Bifidobacterium, Scardovia, Atopobium, Olsenella, Bacteroides, Phocaeicola, Clostridiales vadinBB60 NA, Pseudoramibacter, Parvimonas, Incertae Sedis, Shuttleworthia, Filifactor, Dialister, Megasphaera, Fretibacterium, Alloscardovia, Cryptobacterium, Eubacterium brachy group, Mycoplasma* |

**Appendix Table 2. Characteristics of study participants with different dietary intake clusters defined based on food intake variables.**

| **Characteristics** | **All (n=3254)** | **Dietary Cluster 1 (n=1336)** | **Dietary Cluster 2 (n=1250)** | **Dietary Cluster 3 (n=668)** | ***p* value** |
| --- | --- | --- | --- | --- | --- |
| **Age (yr)** | 48.5 (39.0 - 58.0) | 47.0 (39.0 - 56.0) | 46.0 (37.0 - 56.0) | 53.0 (44.0 - 61.0) | **<0.001** |
| **Sex** |  |  |  |  | **<0.001** |
| Male | 1691 (52.0%) | 806 (60.3%) | 648 (51.8%) | 237 (35.5%) |  |
| Female | 1563 (48.0%) | 530 (39.7%) | 602 (48.2%) | 431 (64.5%) |  |
| **Race** |  |  |  |  | **<0.001** |
| Mexican American | 525 (16.1%) | 151 (11.3%) | 307 (24.6%) | 67 (10.0%) |  |
| Other Hispanic | 314 (9.6%) | 106 (7.9%) | 123 (9.8%) | 85 (12.7%) |  |
| Non-Hispanic White | 1135 (34.9%) | 449 (33.6%) | 492 (39.4%) | 194 (29%) |  |
| Non-Hispanic Black | 864 (26.6%) | 514 (38.5%) | 196 (15.7%) | 154 (23.1%) |  |
| Other Race-Including Multi-Racial | 416 (12.8%) | 116 (8.7%) | 132 (10.6%) | 168 (25.1%) |  |
| **Education Level** |  |  |  |  | **<0.001** |
| Less than 9th grade | 347 (10.7%) | 107 (8.0%) | 178 (14.2%) | 62 (9.3%) |  |
| 9-11th grade | 516 (15.9%) | 249 (18.6%) | 204 (16.3%) | 63 (9.4%) |  |
| High school graduate/GED or equivalent | 715 (22.0%) | 335 (25.1%) | 266 (21.3%) | 114 (17.1%) |  |
| Some college or AA degree | 890 (27.4%) | 390 (29.2%) | 336 (26.9%) | 164 (24.6%) |  |
| College graduate or above | 784 (24.1%) | 253 (18.9%) | 266 (21.3%) | 265 (39.7%) |  |
| Don't know | 2 (0.1%) | 2 (0.1%) | 0 (0.0%) | 0 (0.0%) |  |
| **Income-to-poverty ratio** | 1.9 (1.0 - 4.1) | 1.8 (1.0 - 3.7) | 1.7 (1.0 - 3.8) | 2.9 (1.3 - 5.0) | **<0.001** |
| **BMI (kg/m^2^)** | 28.5 (24.9 - 33.1) | 28.8 (25.1 - 33.5) | 28.7 (25.0 - 33.3) | 27.0 (24.3 - 31.8) | **<0.001** |
| **Tooth count** | 25.0 (21.0 - 28.0) | 25.0 (20.0 - 27.0) | 25.0 (21.0 - 28.0) | 25.0 (22.0 - 28.0) | **<0.001** |
| **Smoking status** |  |  |  |  | **<0.001** |
| Never | 1738 (53.4%) | 613 (45.9%) | 693 (55.4%) | 432 (64.7%) |  |
| Quitted | 734 (22.6%) | 285 (21.3%) | 275 (22.0%) | 174 (26.0%) |  |
| Current smoker: <10/day | 309 (9.5%) | 161 (12.1%) | 113 (9.0%) | 35 (5.2%) |  |
| Current smoker: 10-20/day | 242 (7.4%) | 139 (10.4%) | 87 (7.0%) | 16 (2.4%) |  |
| Current smoker: ≥20/day | 231 (7.1%) | 138 (10.3%) | 82 (6.6%) | 11 (1.6%) |  |
| **Oral conditions** |  |  |  |  | **<0.001** |
| Oral health | 976 (30.0%) | 332 (24.9%) | 369 (29.5%) | 275 (41.2%) |  |
| Caries-only | 420 (12.9%) | 177 (13.2%) | 185 (14.8%) | 58 (8.7%) |  |
| Periodontitis-only | 813 (25.0%) | 326 (24.4%) | 284 (22.7%) | 203 (30.4%) |  |
| Co-existing caries and periodontitis | 1045 (32.1%) | 501 (37.5%) | 412 (33.0%) | 132 (19.8%) |  |
| **Hypertension status** |  |  |  |  | **<0.001** |
| Normal | 1339 (41.1%) | 491 (36.8%) | 554 (44.3%) | 294 (44.0%) |  |
| Elevated | 539 (16.6%) | 225 (16.8%) | 208 (16.6%) | 106 (15.9%) |  |
| Hypertension stage 1 | 747 (23.0%) | 328 (24.6%) | 283 (22.6%) | 136 (20.4%) |  |
| Hypertension stage 2 | 540 (16.6%) | 250 (18.7%) | 175 (14.0%) | 115 (17.2%) |  |
| Hypertension Crisis | 25 (0.8%) | 13 (1.0%) | 7 (0.6%) | 5 (0.7%) |  |
| Missing | 64 (2.0%) | 29 (2.2%) | 23 (1.8%) | 12 (1.8%) |  |
| **Diabetes status** |  |  |  |  | 0.718 |
| Non-diabetes | 1770 (54.4%) | 705 (52.8%) | 700 (56.0%) | 365 (54.6%) |  |
| Prediabetes | 981 (30.1%) | 423 (31.7%) | 356 (28.5%) | 202 (30.2%) |  |
| Diabetes | 235 (7.2%) | 91 (6.8%) | 96 (7.7%) | 48 (7.2%) |  |
| Uncontrolled diabetes | 162 (5.0%) | 73 (5.5%) | 59 (4.7%) | 30 (4.5%) |  |
| Missing | 106 (3.3%) | 44 (3.3%) | 39 (3.1%) | 23 (3.4%) |  |

*Note:*

Oral health: individuals with neither caries and nor periodontitis, using “no periodontitis” based on CDC/AAP definition; Caries: individuals with caries present and with no periodontitis based on CDC/AAP definition; Periodontitis: individuals with no caries and with mild/moderate/severe periodontitis based on CDC/AAP definition; Caries + Periodontitis: individuals with both caries and periodontitis present based on CDC/AAP definition.

BMI (body mass index): calculated as weight (kg) divided by height squared (m^2^).

Income-poverty ratio: calculated by dividing total family income by the poverty threshold based on poverty guidelines, specific to family size, year and state.

Energy intake was assessed as the average of total energy intake derived from two 24-hour dietary recall interviews.

Hypertension status: categorized based on examined systolic and diastolic blood pressure values (SBP and DBP). Normal: SBP <120 mmHg and DBP <80 mmHg; Elevated: SBP 120-129 mmHg and DBP <80 mmHg; hypertension stage I: SBP 130-139 mmHg or DBP 80-89 mmHg; hypertension stage II: SBP ≥140 mmHg or DBP ≥90 mmHg; hypertension crisis: SBP >180 mmHg and/or DBP >120 mmHg.

Diabetes status: defined by tested glycohemoglobin (HbA1c) levels. Non-diabetes: HbA1c <5.7%; Prediabetes: HbA1c 5.7-6.4%; Diabetes: HbA1c ≥6.5%; Poorly controlled diabetes: HbA1c ≥8.0%.

**Appendix Spreadsheet 1. Complete mediation analysis results for dietary indices and representative food intake variables across oral condition outcomes**
